# Impact of health warning labels and calorie labels on selection and purchasing of alcoholic and non-alcoholic drinks: a randomised controlled trial

**DOI:** 10.1101/2022.07.22.22277929

**Authors:** Natasha Clarke, Jennifer Ferrar, Emily Pechey, Minna Ventsel, Mark A Pilling, Marcus R Munafò, Theresa M Marteau, Gareth J Hollands

## Abstract

**Objective:** Health warning and calorie labels on alcohol have the potential to reduce consumption at population level but remain unevaluated using robust designs with behavioural outcomes. The aim of the current study is to estimate the impact on selection and actual purchasing of (a) health warning labels (text-only and image-and-text) on alcoholic drinks and (b) calorie labels on alcoholic and non-alcoholic drinks.

**Design:** Parallel-groups randomised controlled trial.

**Setting:** Participants selected drinks in a simulated online supermarket, before purchasing them in an actual online supermarket.

**Participants:** Adults in England and Wales who regularly consumed and purchased beer or wine online (n= 644).

**Interventions:** Participants were randomised to one of six groups in a between-subjects 3 (*Health warning labels (HWLs)*: i. image-and-text HWL, ii. text-only HWL, iii. no HWL) x 2 (*Calorie labels*: present vs absent) factorial design.

**Main outcome measures:** The number of alcohol units selected (with intention to purchase); secondary outcomes included alcohol units purchased and calories selected and purchased.

**Results:** 608 participants completed the study and were included in the primary analysis. There was no evidence of an overall difference for either (a) HWLs, or (b) calorie labels on the number of alcohol units selected [HWLs: F(2,599) = 0.406, p = .666; calorie labels: F(1,599) = 0.002, p = .961]. There was also no evidence of an overall difference on any secondary outcomes, including the number of alcohol units purchased [HWLs: F(2,462) = 1.85, p = .159; calorie labels: F(1,462) = 0.193, p = .661].

In pre-specified subgroup analyses comparing the ‘calorie label only’ group (n = 101) to the ‘no label’ group (n = 104) there was no evidence that calorie labels reduced the number of calories selected [unadjusted means: 1913 calories vs 2203 calories, p = .643]. Amongst the 75% of participants who went on to purchase drinks, those in the ‘calorie label only’ group (n = 74) purchased fewer calories than those in the ‘no label’ group (n = 79) [unadjusted means: 1532 calories vs 2090, p = .028].

**Conclusion:** There was no evidence that health warning labels reduced the number of alcohol units selected or purchased in an online purchasing setting. There was some evidence suggesting that calorie labels on alcoholic and non-alcoholic drinks may reduce calories purchased. Further evaluation is warranted in suitably powered studies in real world settings.

**Trial registration:** Pre-registered protocol (https://osf.io/ch2sm/) and prospective ISCRTN registration: https://www.isrctn.com/ISRCTN10313219

**Funding:** This study was funded by Wellcome [Grant number 206853/Z/17/Z].

## Introduction

Excessive alcohol consumption is a major contributor to the global burden of non-communicable diseases, such as cancer, heart disease and stroke ^1,2^. Interventions that alter the physical and economic environments in which alcohol-related behaviours occur have the potential to reduce its consumption ^3^. Improved labelling of alcohol products is one intervention that has been proposed, with potential to be implemented at scale ^4,5^.

There is strong evidence that tobacco health warning labels (HWLs) increase a range of smoking cessation-related behaviours ^6,7^, and are a feasible population level intervention ^8^. In addition, these effects are evident in similar magnitude amongst those in more and less deprived groups ^9^. Evidence from online studies suggests that while both image-and-text HWLs – which include an often aversive visual image -and text-only HWLs, reduce hypothetical selection of alcoholic drinks, the former are more effective ^10^. Initial laboratory studies suggest both are similarly effective at decreasing consumption rate ^11^ but that image-and-text HWLs may exert larger effects on abstinence and consumption intentions than text-only HWLs ^12^. There is an absence of evidence of the impact of HWLs from field settings, such as online and physical supermarkets ^13,14^. While a previous study in a naturalistic shopping laboratory found no impact of HWLs on selection or purchasing behaviour ^15^, the setting lacked ecological validity as no money was exchanged and participants did not keep the drinks they selected.

Another potential labelling intervention is the provision of calorie information, which current evidence suggests may have small effects on healthier selection and consumption of food products ^16^. Many alcohol products are currently exempt from mandatory nutrition labelling, including in the UK Government’s recently implemented policy (April 2022) on calorie labelling out of the home ^17^. This is despite alcohol being the second most energy dense foodstuff (7.1kcal/g) after fat (9kcal/g). Most products therefore do not display this information ^18^ and as a result drinkers’ knowledge of the energy content of alcoholic drinks is poor ^19^. However, the UK Government’s most recent obesity strategy included plans to consult on the provision of calories on alcohol ^20^ and there are increasing calls for improved alcohol labelling, including through displaying calorie information ^5,21^. Current evidence on the provision of calorie information on alcohol is scarce, with a recent review finding no studies in real-world settings or of effects on actual purchasing behaviour ^22^.

The principal aim of this study was to estimate the impact on selection of (a) HWLs (text-only and image-and-text) on alcoholic drinks and (b) calorie labels on alcoholic and non-alcoholic drinks. It was hypothesised that HWLs and calories labels would reduce the number of alcohol units selected.

## Methods

The study was prospectively registered (https://www.isrctn.com/ISRCTN10313219). In addition, both the study protocol (https://osf.io/ch2sm/) and a detailed statistical analysis plan (https://osf.io/qwdra/) were pre-registered on the Open Science Framework (OSF). The study was approved by the University of Cambridge ethics committee (ref: PRE.2020.155). Trial reporting follows CONSORT 2010 guidelines.

### Study design

The study used a parallel-groups 3 (*Health warning labels (HWLs)*: i. image-and-text HWL, ii. text-only HWL, iii. no HWL) x 2 (*Calorie labels*: present vs absent) factorial design (see Box 1).

**Box 1.**
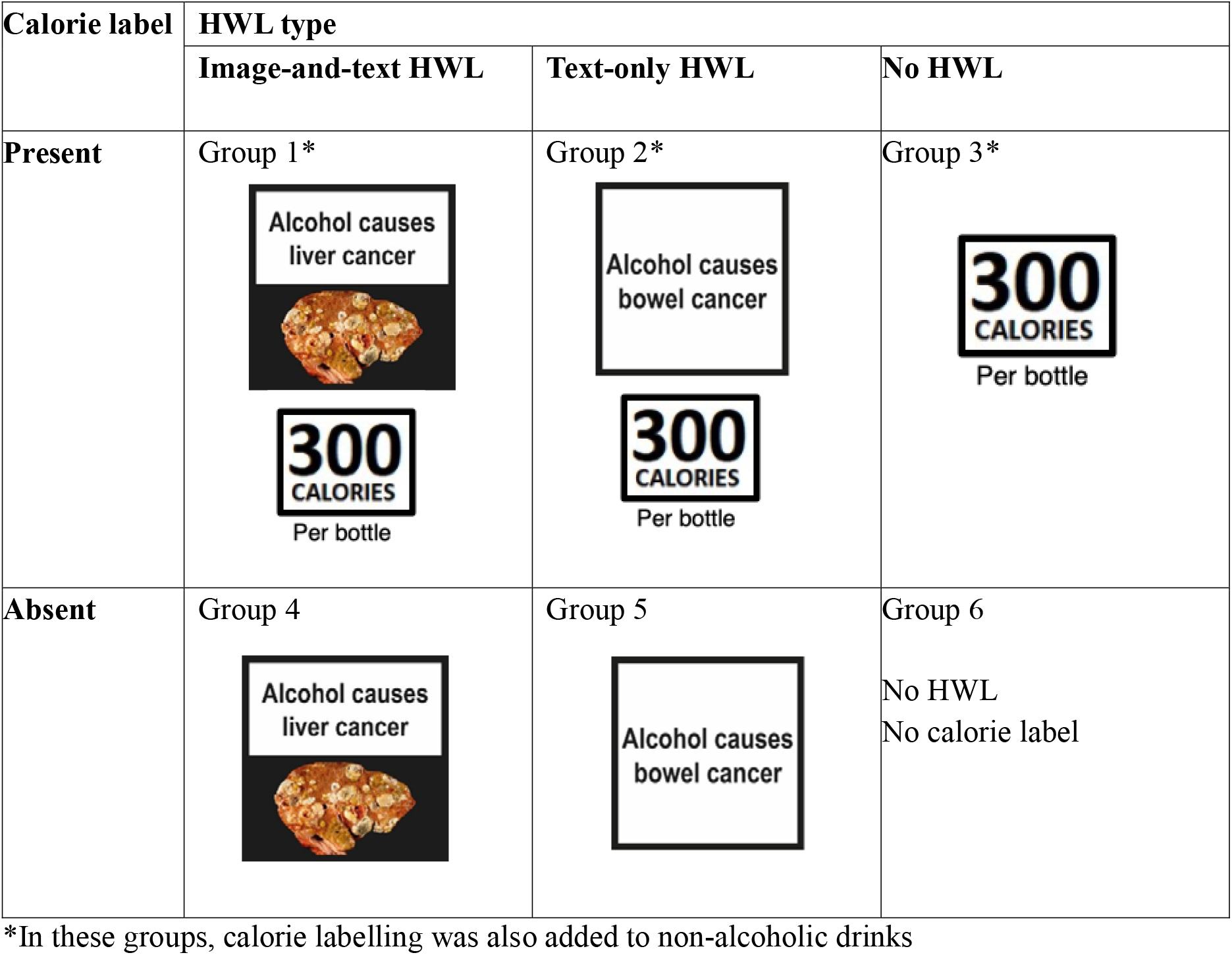
Health warning label (HWL) type and calorie label displayed with the alcoholic drinks in the selection task.

### Setting

The study was conducted online using simulated and real online supermarkets. First, participants completed a simulated supermarket selection task hosted on the Qualtrics online survey platform (see Figure 1). Following this, participants were required to purchase the same drinks in Tesco online supermarket (Tesco.com), the largest national supermarket in the UK.

**Figure 1.**
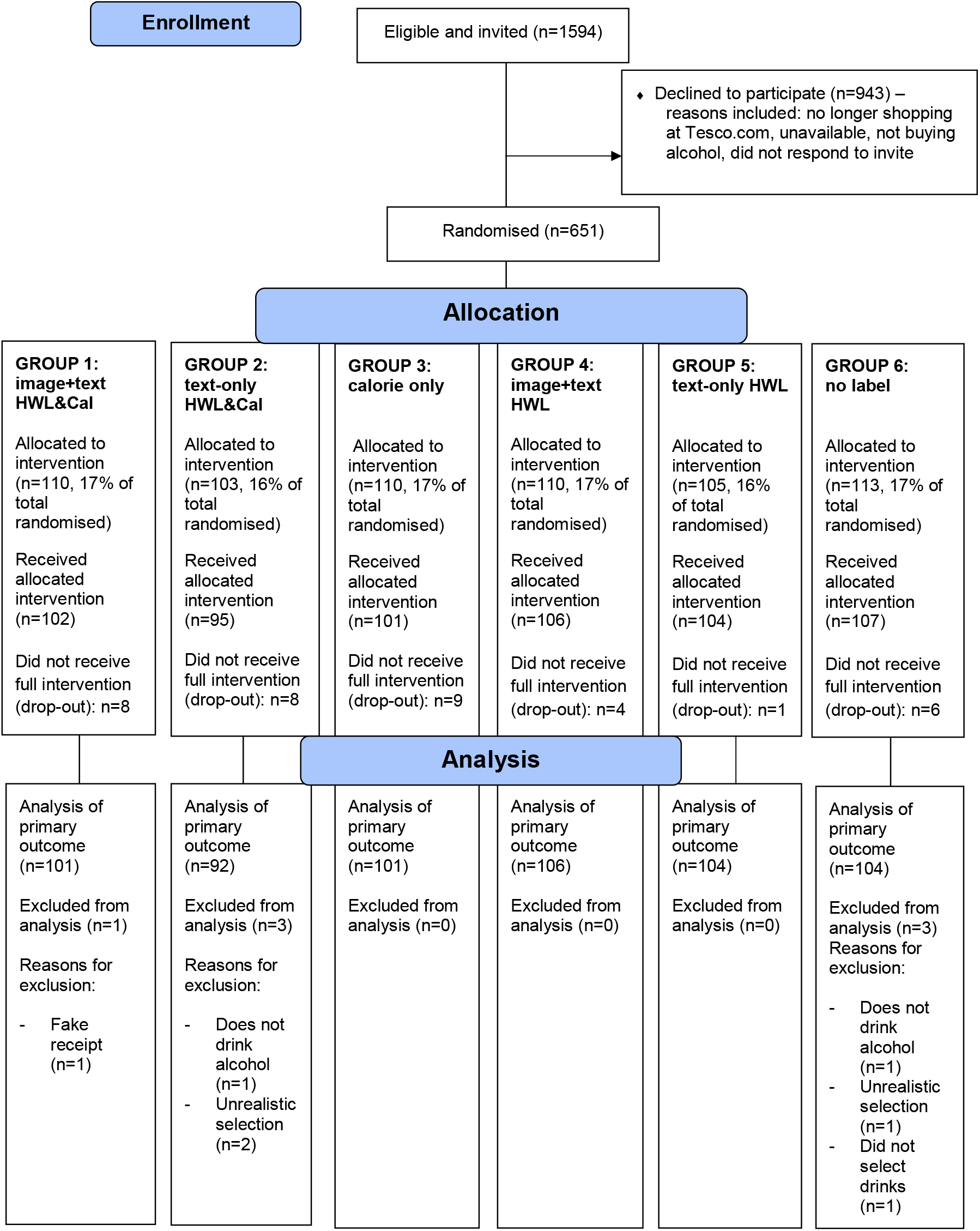
Flow of participants through study

### Participants

Eligible participants were adults (18+) residing in England or Wales, who self-reported that they consumed beer or wine regularly (i.e., at least weekly), and purchased these drinks at least monthly from Tesco.com, with a minimum spend of £20. Participants had to be willing to complete a shop at Tesco.com following completion of the selection task, book a delivery or click-and-collect slot, and send proof of purchase (their receipt) to the research team. Similar proportions of males and females of a range of ages were recruited via Roots Research (https://rootsresearch.co.uk/), one of the largest research agencies in the UK, with a high-quality panel of over 350,000 participants. Recruitment occurred between September 2021-March 2022.

#### Sample size

There was no direct evidence available within the literature from which to estimate the effect of the intervention on selection of multiple drink options. There was also no prior information on the size of interaction effect for HWLs and calorie labels. A maximum sample size of 600 was possible with available resources (100/per group). A similar online study of alcohol HWLs ^10^ found the proportion of participants selecting an alcoholic drink was lower when alcohol products had an image-and-text HWL (56%) or text-only HWL (61%), compared to no HWLs (77%) (i.e., a 16-21% point difference). However, only a single drink was selected in this online study and there was no intention to purchase the selected drinks, or opportunity to do so. An illustrative sample size calculation based on 600 participants suggests that with 85/group (allowing for attrition of 15%) there would be 80% power and at alpha 5% (510 participants) to detect an overall interaction effect size f of 0.147 or greater with a 2-way ANOVA.

### Randomisation and masking

Randomised assignment of participants was completed via the default algorithm in Qualtrics with a ratio of 1:1:1:1:1:1. Participants were unaware of their group assignment throughout the study. The research team were blinded to allocation until participants had completed the primary outcome and there was no possibility of contact between the research team and participants until after the primary outcome and the selection task were completed; the statistician completing the analysis was blinded to the allocation.

### Intervention

All participants viewed a total range of 64 drink options. This comprised i) a range of beers, ciders, alcohol-free beer and cider alternatives, and soft drinks (32 options), and ii) a range of wines, alcohol-free wine alternatives, and soft drinks (32 options), modelled on the available range of products on Tesco.com. Alcoholic drinks were labelled according to the six groups: 1. image-and-text HWL and calorie label, 2. text-only HWL and calorie label, 3. calorie calorie label only, 4. image-and-text HWL only, 5. text-only HWL only, 6. no HWL or calorie label (Box 1). In the calorie label groups (1,2,3), non-alcoholic drinks also displayed calorie labels. To ensure they were clearly visible, labels were displayed next to the product. The specific warnings used were developed and tested in previous studies and were HWLs that were most effective in increasing negative emotions ^23^ and decreasing the odds of selecting alcohol ^10^. Within the HWL groups, eight different variants of image-and-text HWLs and seven different variants of text-only HWLs were used to increase variety, maximise engagement and likelihood of impact across our sample. This is also in line with tobacco guidelines specifying rotating warnings are most effective ^24^. Calorie information was given per bottle or can if the total volume of the container was 568ml or less, or by glass (250ml) if the volume was over 568ml (i.e., 1 pint or less). Illustrative examples of labelled alcohol products and the full drinks list are included in Supplementary Material S1.

Price promotions and variations for all drinks included in the selection task were checked every month via Tesco.com and recorded. Prices shown in the task reflected the full price on Tesco.com throughout the study and did not reflect offers or promotions. If there was a 20% or more price change during the study, then the price for that product was updated. Participants were informed that the prices reflect Tesco prices on the date the study started and that there might be slight variations.

In the Typology of Interventions in Proximal Physical Micro□Environments (TIPPME) ^3^, both health warning and calorie labelling interventions are classified as *‘Information x Product’* interventions.

### Outcome measures

#### Primary outcome

The primary outcome was the number of alcohol units selected in the context of a stated intention to purchase. A unit is a standard measure of pure alcohol in a drink and in the United Kingdom one unit is 10ml or 8g of pure alcohol. Participants were aware when selecting drinks in the task that they were required to subsequently purchase the drinks chosen and send proof of this to the research team (otherwise they were not reimbursed). Units of alcohol were calculated for all drinks that were >0% ABV, i.e., alcoholic and ‘alcohol-free’ drinks (which were defined as containing between 0% and up to 0.5% ABV). This outcome was pre-registered as the primary outcome as it was assessed in all participants who were exposed to the intervention, and measured within the same context, i.e., the simulated online supermarket.

### Secondary outcome measures

Secondary outcomes were the number of alcoholic drinks selected; the number of non-alcoholic drinks selected; the number of alcohol units purchased; the proportion (i.e., percentage) of total drinks selected that were alcoholic; the proportion (i.e., percentage) of total drinks purchased that were alcoholic; the total number of calories selected (overall and by drink category: alcoholic and non-alcoholic drinks); the total number of calories purchased (overall and by drink category: alcoholic and non-alcoholic drinks).

Additional outcomes were the total number of drinks selected; the total number of drinks purchased; the number of alcoholic drinks purchased; the number of non-alcoholic drinks purchased.

Selection outcomes were assessed from the simulated online supermarket task and purchasing outcomes were assessed from receipts after shops at Tesco.com were completed.

### Other measures

#### Negative emotional arousal generated by health warning labels

was assessed using a four-item measure, previously used to assess the impact of warning labels on cigarette packages ^25^ and adapted for alcohol HWL studies ^10,15^. Responses were rated on seven point scales: ‘How [afraid/worried/uncomfortable/disgusted] does the label on this drink make you feel?’ (1 Not at all [afraid/worried/uncomfortable/disgusted] to 7 very [afraid/worried/uncomfortable/disgusted]).

#### Acceptability of health warning labels

was assessed using one item on a 7-point scale, adapted from previous research assessing the impact of sugar tax ^26^ and alcohol HWLs ^10,15^. ‘Do you support or oppose putting this label on alcoholic drinks?’ (Strongly oppose–neither oppose nor support–strongly support). Ratings past the scale midpoint (indicating neither acceptable nor unacceptable), *i*.*e*., >4, indicated that the label was acceptable.

#### Demographic characteristics

Age, gender, and highest qualification attained (‘Higher Education or professional / vocational equivalents’, ‘A levels or vocational level 3 or equivalents’, ‘GCSE / O Level grade A*□C or vocational level 2 or equivalents’, ‘Qualifications at level 1 and below’, ‘Other qualifications: level unknown’, or ‘No□qualifications’). Qualifications classifications were based on UK definitions ^27,28^.

#### Household members

Number of adults (aged 18+) and of children (aged <18).

#### Drinking behaviour risk

Alcohol Use Disorders Identification Test (AUDIT) ^29^, a 10-item clinical screening measure for assessing risk associated with participants’ drinking behaviour (low risk drinking: score 0-7; medium/hazardous risk drinking: score 8-15; high/harmful risk drinking: score ≥16).

#### Baseline weekly unit consumption

Self-reported drinks consumed and purchased over the previous seven days, used to calculate the number of alcohol units as a continuous variable.

#### Manipulation check

Participants were asked if they noticed any labels on the products and to describe these. They were also asked what they thought the study was about.

#### Open text comments

Participants provided comments on the task, such as explaining their choice of drinks.

### Procedure

Participants were initially provided with an information sheet, instructions, and a link to the study via email. Participants were told the study was investigating “Drink choices and shopping behaviour” and were not made aware of the study aim. At the start of the study task, participants were again presented with this information and provided informed consent. Participants were randomised and in a simulated online supermarket environment replicating Tesco.com (presented within Qualtrics) they were shown the available drink selection. They chose all the drinks they wanted to purchase in their next online shop at Tesco.com. They were then shown their total drink selection and price, and given the opportunity to amend their selection before continuing. Participants then completed demographic and drinking behaviour measures.

After completing the simulated online supermarket task, participants were automatically sent an email detailing their selection, alongside further instructions for completing purchasing of the same items, including a direct link to Tesco.com. Participants were instructed to immediately place their selected drinks in their Tesco.com shopping basket, along with any other items, then book their delivery or collection slot, and confirm this within 48 hours. They were sent a reminder email on their delivery/collection day and requested to send an itemised receipt to the research team within 48 hours. Up to two follow-up reminders were sent, two and four days later. Purchases were recorded from receipts, including any additional drink purchases. Substitutions by the participant or by Tesco that were explained (e.g., not in stock) were marked as the original drink they attempted to purchase. Participants were debriefed via email and reimbursed £35(∼$44) for their time taking part in the study (but not for the drinks they purchased).

### Statistical analysis

Analyses were pre-registered in a detailed statistical analysis plan (https://osf.io/qwdra/).

All participants who completed the selection task were included in the primary outcome analysis. Participants who failed to complete the selection task and those whose responses were flagged as incomplete or suspicious - e.g., those that forged data (i.e., submitted fake receipts that were not generated by Tesco) or selected an unrealistically large number of drinks (e.g., over 200 units) that were not purchased – were excluded (see Figure 1 for details by group).

For the primary outcome a generalised linear model was used. An overall 2-way ANOVA summary and the equivalent regression summary are reported (see Supplementary Material S2a). The model utilised the 3 × 2 design with two independent variables: 1) image-and-text HWL vs. text-only HWL vs. no label, and 2) calorie vs. no calorie labelling. Umbrella ANOVA p-values, at a threshold for significance of 0.025 (i.e., 5%/2) are reported. The interaction terms were dropped as there was no clear evidence of an interaction (p > 0.01).

For most secondary and additional outcomes, ANCOVA and regression models were repeated as per the primary outcome model. For calories selected from non-alcoholic drinks and calories purchased from alcoholic and non-alcoholic drinks the distribution was highly skewed and zero inflated, and therefore a hurdle model was used for analysis, fitting i) a binary logistic model (part 1) to the zero and non-zero outcomes and ii) a truncated negative binomial model (part 2) to just the positive values. The regression model results for the positive values are therefore based only on participants who selected at least one drink or one drink containing calories (see Supplementary Material S2b), with non-integer variables rounded to integer values before hurdle model analysis. For total calories selected and total alcohol units purchased outcomes, negative binomial regression was required due to the skewed data. For the proportion outcomes (i.e., percentage of total drinks selected, and purchased, that were alcoholic), a beta binomial regression was used to model the proportion using the counts of relevant drinks selected out of the count of all drinks selected as this could accommodate the bimodal distribution observed. For these outcomes only, due to the nature of the model, any participants who did not select any drink (as appropriate for the outcome) were excluded. For calorie outcomes, only the main effect of calorie labelling is reported from the model, with HWLs interpreted as a covariate. The calorie outcome analyses also prioritised the reporting of 1/15 of the pairwise comparisons, for the one condition level testing the contrast between calorie labelling (without HWLs) and no calorie labelling (without HWLs). For label ratings (negative emotional arousal and acceptability), a generalised linear model compared the labelling groups in pairwise comparisons, including the original group membership as a covariate.

Two per-protocol analyses were pre-specified each for the alcohol units and calories outcome, in which the primary outcome analysis was repeated for (i) participants who purchased what they selected, either with or without additional drinks (per-protocol analysis 1); (ii) only participants who purchased exactly what they selected and purchased with no additional drinks (per-protocol analysis 2).

Open text comments provided by participants were manually coded and emergent themes were identified. Themes were agreed between all authors. For full details of the analytic procedure see Supplementary Material S3.

### Patient and public involvement

The design and implementation of the study, including the plans for recruitment and measurement of the outcomes were independent of patients and the public. Patients or members of the public were not invited to comment on the study design or contribute to the writing or editing of this document for readability or accuracy. The results of the research will be shared with the general public through internet, news, and popular science articles and social media.

## Results

### Sample characteristics

Figure 1 shows the flow of participants. In total, 651 participants were randomised, 615 of whom completed the selection task. 608 participants were included in the primary outcome analysis. For purchasing outcomes, of the 608 participants who completed the selection task, 467 (77%) went on to purchase drinks from Tesco.com (this was similar across groups: range 73% to 80%). The primary analysis sample was 55% female, and the mean age was 35.5 (SD = 10.8). Groups were well balanced on most characteristics (Table 1). The number of units consumed and purchased was lower in Group 3 (calorie only label group). Weekly units purchased was therefore included as a covariate in the models.

**Table 1.** Characteristics of participants included in the primary outcome analysis.

|  | Calorie label |  |  |  |  |  |
| --- | --- | --- | --- | --- | --- | --- |
|  | Present |  |  | Absent |  |  |
|  | **Group 1:**<br>**Image-and-text**<br>**HWL**<br>**n = 101** | **Group 2:**<br>**Text-only HWL**<br>**n = 92** | **Group 3:**<br>**No HWL**<br>**n = 101** | **Group 4:**<br>**Image-and-text**<br>**HWL**<br>**n = 106** | **Group 5:**<br>**Text-only HWL**<br>**n = 104** | **Group 6:**<br>**No label**<br>**n = 104** |
| **Alcohol consumption previous week (units) (mean (SD))** | 24.86 (24.38) | 23.80 (22.36) | 19.03 (18.13) | 25.65 (22.80) | 22.31 (21.85) | 29.62 (32.25) |
| **Alcohol purchasing previous week (units) (mean (SD))** | 34.07 (29.25) | 30.58 (27.52) | 25.35 (22.72) | 33.68 (27.05) | 31.32 (21.78) | 33.75 (28.36) |
| **AUDIT score (mean (SD))** | 8.8 (4.3) | 9.62 (5.37) | 8.06 (4.35) | 9.78 (5.32) | 10.32 (5.59) | 9.76 (5.52) |
| - Low risk drinking (scores 1-7) | 45 (45) | 36 (40) | 50 (50) | 37 (35) | 37 (36) | 43 (41) |
| - Medium to high risk drinking scores (8 +) | 56 (55) | 56 (60) | 51 (50) | 69 (65) | 67 (64) | 61 (59) |
| **Age (mean (SD))** | 35.64 (10.96) | 33.66 (10.39) | 35.53 (10.51) | 35.93 (10.21) | 35.43 (11.60) | 36.79 (10.90) |
| 18-39 years | 28 (28) | 19 (21) | 26 (26) | 32 (30) | 30 (29) | 35 (34) |
| 40 and over | 73 (72) | 73 (79) | 75 (74) | 74 (70) | 74 (71) | 69 (66) |
| **BMI (mean (SD))** | 26.90 (5.46) | 25.71 (5.73) | 25.43 (5.01) | 26.65 (5.74) | 26.56 (6.07) | 25.92 (5.31) |
| **Gender (n (%))** |  |  |  |  |  |  |
| Male | 39 (38.6) | 44 (47.8) | 37 (36.6) | 52 (49.1) | 47 (45.2) | 50 (48.1) |
| Female | 61 (60.4) | 48 (52.2) | 63 (62.4) | 54 (50.9) | 57 (54.8) | 54 (51.9) |
| Other |  |  |  |  |  |  |
| **Household members** |  |  |  |  |  |  |
| Number of adults in household (mean (SD)) | 1.87 (0.82) | 2.22 (2.11) | 1.93 (0.67) | 2.08 (0.70) | 2.07 (0.95) | 1.96 (0.72) |
| Number of children in household (mean (SD)) | 0.70 (0.97) | 0.64 (0.99) | 0.67 (0.98) | 0.81 (0.99) | 0.67 (0.92) | 0.81 (1.0) |
| **Highest qualification (n (%))** |  |  |  |  |  |  |
| No qualifications | 1 (1) | 0 (0) | 1 (1) | 2 (2) | 0 (0) | 0 (0) |
| Qualifications at level 1 and below | 0 (0) | 1 (1) | 0 (0) | 1 (1) | 0 (0) | 1 (1) |
| GCSE / O Level grade A*–C or vocational level 2 or equivalents | 11 (11) | 11 (12) | 18 (18) | 10 (9) | 11 (11) | 14 (13) |
| A levels or vocational level 3 or equivalents | 19 (19) | 16 (17) | 17 (18) | 18 (17) | 21 (20) | 25 (24) |
| Higher Education or professional / vocational equivalents | 69 (68) | 64 (70) | 64 (63) | 74 (70) | 71 (68) | 64 (62) |
| Other qualification | 1 (1) | 0 (0) | 0 (0) | 1 (1) | 1 (1) | 0 (0) |
| **Ethnicity (n (%))** |  |  |  |  |  |  |
| White British / White Irish / Other White background | 81 (80) | 67 (73) | 76 (75) | 86 (81) | 83 (80) | 83 (80) |
| Mixed White and Black African / Mixed White and Asian / Mixed White and Black Caribbean / Other Mixed background | 6 (6) | 2 (3) | 5 (5) | 3 (3) | 7 (7) | 5 (5) |
| Asian/Asian British (Indian/Pakistani/Bangladeshi/Chinese/Other Asian/Asian British background) | 9 (9) | 17 (19) | 12 (12) | 9 (8) | 5 (5) | 10 (10) |
| Black/Black British (African/Caribbean/Other Black/Black British background) | 5 (5) | 4 (4) | 7 (7) | 7 (7) | 7 (6) | 3 (3) |
| Other ethnic group | 0 (0) | 1 (1) | 0 (0) | 0 (0) | 2 (2) | 2 (2) |
| *Prefer not to say/missing* | 0 (0) | (0) | 1 (1) | 1 (1) | 0 (0) | 1 (1) |

Raw primary and secondary outcome data are in Table 2 and model results in Table 3.

**Table 2.** Primary and secondary outcomes: raw means (+/-SDs), by group.

|  | Calorie label |  |  |  |  |  |
| --- | --- | --- | --- | --- | --- | --- |
|  | Present |  |  | Absent |  |  |
|  | Group 1:<br>Image-and-text HWL<br>n = 101 | Group 2:<br>Text-only HWL<br>n = 92 | Group 3:<br>No HWL<br>n = 101 | Group 4:<br>Image-and-text HWL<br>n = 106 | Group 5:<br>Text-only HWL<br>n = 104 | Group 6:<br>No label<br>n = 104 |
| **Primary outcome** |  |  |  |  |  |  |
| Total alcohol units selected | 25.7 (24.4) | 22 (18.8) | 19.6 (20.9) | 21.3 (16.3) | 22.0 (16) | 23.7 (17.5) |
| **Secondary outcomes: selection** |  |  |  |  |  |  |
| *Alcohol* |  |  |  |  |  |  |
| Number of alcoholic drinks selected | 9.38 (9.95) | 7.24 (8.02) | 6.58 (10.4) | 7.43 (8.27) | 7.61 (7.83) | 7.64 (8.02) |
| Number of non-alcoholic drinks selected | 6.64 (9.64) | 7.76 (16.5) | 5.99 (9.77) | 5.34 (7.28) | 4.63 (6.51) | 5.77 (8.23) |
| Proportion of alcoholic drinks selected that are alcoholic | 0.64 (0.35) | 0.57 (0.38) | 0.55 (0.36) | 0.59 (0.38) | 0.63 (0.34) | 0.64 (0.37) |
| *Calories* |  |  |  |  |  |  |
| Total calories selected | 2396 (1943) | 2275 (2016) | 1913 (2097) | 2038 (1386) | 1995 (1509) | 2203 (1589) |
| Calories selected from non-alcoholic drinks | 492 (694) | 657 (1231) | 487 (768) | 447 (614) | 357 (530) | 477 (709) |
| Calories selected from alcoholic drinks | 1903 (1783) | 1618 (1398) | 1427 (1595) | 1592 (1282) | 1638 (1273) | 1725 (1298) |
| **Secondary outcomes: purchasing** |  |  |  |  |  |  |
|  | **n = 74** | **n = 71** | **n = 74** | **n = 85** | **n = 84** | **n = 79** |
| *Alcohol* |  |  |  |  |  |  |
| Total alcohol units purchased | 25.4 (25.9) | 22.8 (18.9) | 17.5 (12.0) | 22.8 (15.5) | 22.2 (15.6) | 22.9 (15.8) |
| Proportion of alcoholic drinks purchased that are alcoholic | 0.64 (0.36) | 0.59 (0.37) | 0.58 (0.36) | 0.64 (0.35) | 0.63 (0.343) | 0.62 (0.38) |
| *Calories* |  |  |  |  |  |  |
| Total calories purchased | 2292 (1846) | 2210 (1652) | 1532 (851) | 2095 (1293) | 1974 (1418) | 2090 (1295) |
| Calories purchased from non-alcoholic drinks | 468 (614) | 509 (685) | 311 (235) | 436 (646) | 328 (501) | 441 (664) |
| Calories purchased from alcoholic drinks | 1874 (1748) | 1701 (1408) | 1221 (878) | 1659 (1152) | 1646 (1239) | 1649 (1150) |

**Table 3.** Model results ANCOVA: F value, degrees of freedom, p values (interaction term not included as p> 0.01)

|  | **HWL (overall) main effect** | **Calorie labelling main effect** |
| --- | --- | --- |
| **Primary outcome** |  |  |
| Total alcohol units selected | F (2,599) = 0.406, p = .666 | F (1,599) = 0.624, p = .430 |
| **Secondary outcomes: selection** |  |  |
| *Alcohol* |  |  |
| Number of alcoholic drinks selected | F (2,599) = 1.042, p = .354 | F (1,599) = 0.063, p = .802 |
| Number of non-alcoholic drinks selected | F (2,599) = 0.016, p = .984 | F (1,599) = 3.482, p = .063 |
| *Calories* |  |  |
| Total calories selected | --- | F (1,599) = 0.624, p = .430 |
| Calories selected from alcoholic drinks | -- | F (1,599) = 0.001, p = .980 |
| Calories selected from non-alcoholic drinks | -- | F (1,599) = 3.23, p = .072 |
| **Secondary outcomes: purchasing** |  |  |
| *Alcohol* |  |  |
| Total alcohol units purchased | F (2,462) = 1.85, p = .159 | F (1, 462) = 0.193, p = .661 |
| *Calories* |  |  |
| Total calories purchased | -- | F (1, 462) = 0.089, p = .766 |
| Calories purchased from alcoholic drinks | -- | F (1, 462) = 0.328, p = .567 |
| Calories purchased from non-alcoholic drinks | -- | F (1, 462) = 0.262, p = .609 |

*N.B. beta-binomial regression models were used for proportion outcomes, these are reported in the regression table, along with the other regression results (Supplementary Material S2a)*

### Primary outcome

There was no evidence of an overall difference for (a) HWLs or (b) calorie labels on the number of alcohol units selected [HWLs: F(2,599) = 0.406, p = .666; calorie labels: F(1,599) = 0.002, p = .961].

### Secondary outcomes

#### Alcohol selected and purchased

There was no evidence of an overall difference for alcohol selection or purchasing on any of the secondary outcomes, including the number of alcohol units purchased [HWLs: F(2,465) = 1.837, p = .16; calorie labels: F(1,465) = 0.22, p = .639].

#### Calories selected and purchased

There was no evidence of an effect of calorie labels on total calories selected [calorie labels: F(1,599) = 0.624, p = .430] or total calories purchased [calorie labels: F(1,462) = 0.089, p = .766].

#### Subgroup analyses of ‘Calorie label only’ group vs ‘No label’ group

These pre-specified analyses compared the effect on calories selected and purchased of those randomised to the ‘calorie label only’ group (Group 3; n = 101) with those randomised to the ‘no label’ group (Group 6; n = 104) (i.e., this analysis included 205 of 608 randomised participants).

There was no evidence of a difference in calories selected between the ‘calorie label only’ and the ‘no label’ groups (p = .643).

Amongst the 75% (153/205 participants) in these two groups who went on to purchase drinks (calorie label only: 74/101; no label: 79/104), fewer calories were purchased by those in the calorie label only group [p = .0282, a reduction of -20% (95%CI: -2%, -35%)]. This effect was evident both for alcoholic [p = .0229, a reduction of 22% (95%CI: -37.18%, -3.41%)] and for non-alcoholic drinks [p = .0086, a reduction of -34% (95%CI: -51.64%, -10.04%)] (see Supplementary Material S2a).

To explore the difference in findings between calorie selection and purchasing, demographic and drinking characteristics of participants that did not go on to purchase (n = 141) were compared to those that did go on to purchase the drinks they selected (n = 467). Purchasers tended to be more educated, older, and self-reported purchasing and consuming less alcohol (see Supplementary Material S4).

### Per-protocol analyses (Table 4)

#### Alcohol selected

There was no evidence for an overall difference of (a) HWLs or (b) calorie labels on the number of alcohol units selected in those that purchased the drinks they selected, either with or without additional drinks (ps > .3).

**Table 4.** Per-protocol analyses: Model results ANCOVA: F value, degrees of freedom, p values.

|  | HWL (overall) main effect | Calorie main effect |
| --- | --- | --- |
| **Alcohol selection** |  |  |
| Per-protocol analysis 1: number of alcohol units selected (exact match, with or without additional drinks) (n=456) | F (2,451) = 0.793, p = .453 | F (1,451) = 1.059, p = .304 |
| Per-protocol analysis 2: number of alcohol units selected (exact match with no additional drinks) (n=426) | F (2,422) = 0.920, p = .399 | F (1,422) = 0.687, p = .408 |
| **Calorie selection** |  |  |
| Per-protocol analysis 1: total calories selected (including all additional drinks) (n=456) | -- | F (1,451) = 0.804, p = .370 |
| Per-protocol analysis 2: total calories selected (exact match with no additional drinks) (n=426) | -- | F (1,422) = 0.417, p = .519 |

#### Calories selected

There was no evidence for an overall difference of calorie labels on the number of calories selected in those that purchased the drinks they selected, either with or without additional drinks (ps > .3).

There was a reduction in number of calories selected in the ‘calorie label only’ group (Group 3) compared to the ‘no label’ group (Group 6) in those that purchased the drinks they selected (n = 205 of 608 randomised), both with additional drinks [p = .024, a reduction of 19% (95% CI: -34%, -1%)] and without [p = .03, a reduction of 20% (95% CI: -35%, -2%)].

### Additional outcomes

There was evidence to suggest image-and-text and text-only HWLs increased the total number of alcoholic drinks purchased [Image-and-text HWL: 42% increase (95% CI: 5%, 80%, p = .03); Text-only HWL: 39% increase (95% CI: 1%, 76%, p = .046)]. There was no evidence for an overall difference of (a) HWLs or (b) calorie labels on any other additional selection or purchasing outcomes (see Supplementary Material S5).

#### Label ratings

Based on comparing means and 95%CIs, calorie labels were rated as more acceptable and had lower scores for negative emotional arousal (i.e., lower fear, disgust, worry, discomfort) than all image-and-text and text-only HWLs. Overall, text-only HWLs were rated as more acceptable and had lower scores for negative emotional arousal than image-and-text HWLs. Negative emotional arousal scores were lower when calorie labels were displayed compared to when there was no calorie label (see Supplementary Material S6).

### Analysis of free-text comments

Four hundred comments were left at the end of the study that contained content suitable for analysis. Attitudes towards calorie labels were generally more positive than to HWLs, with 54% of calorie label-specific comments reflecting positive responses while 24% of HWL-specific comments showed positive responses to HWLs. Responses to HWLs have been qualitatively analysed previous research ^15,23^, but calorie labels have received less attention. Box 2 therefore outlines the main themes that emerged from responses to calorie labels; a detailed overview of themes related to both calorie labels and HWLs can be found in Supplementary Material S3.

**Box 2.**
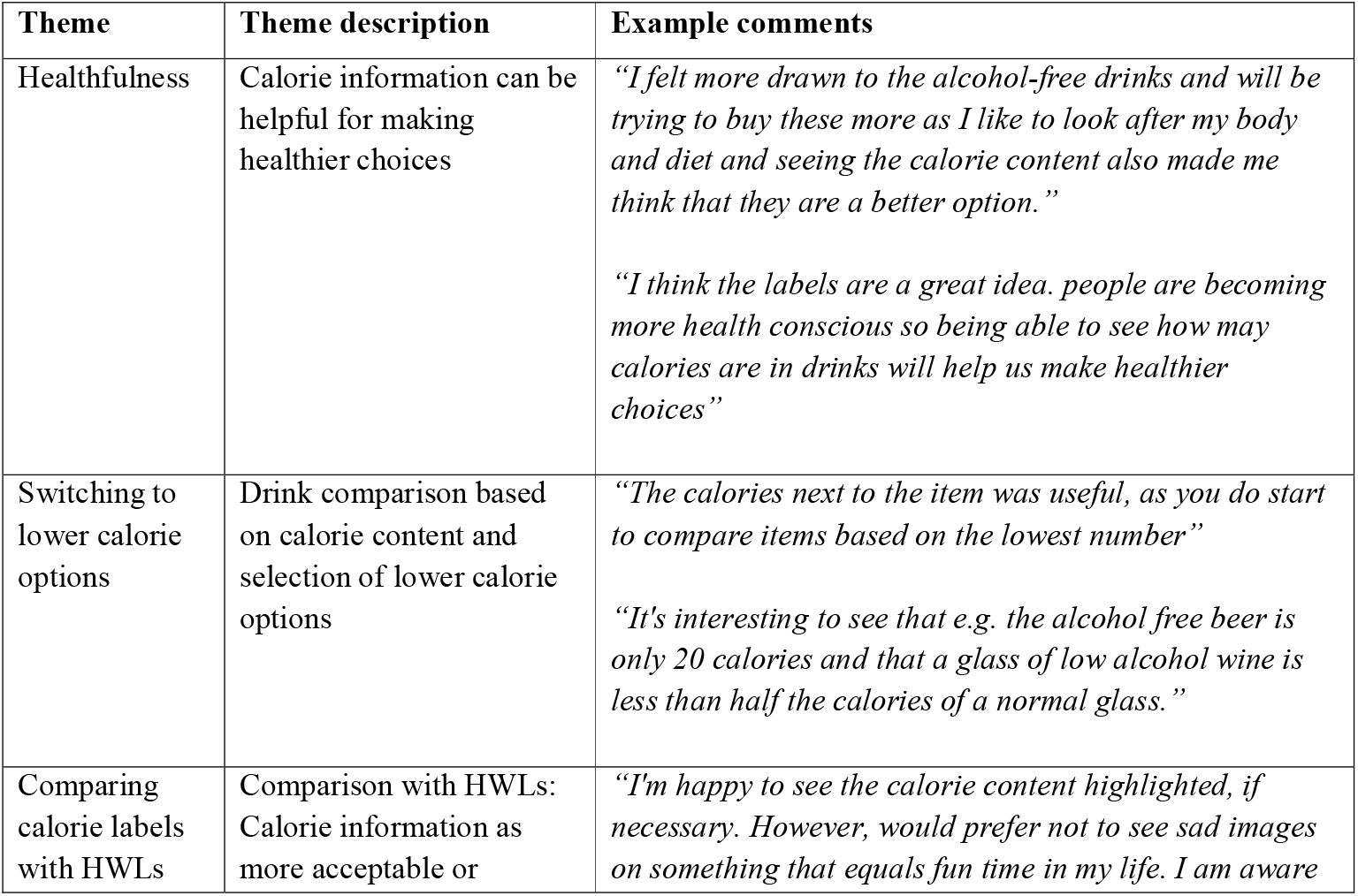
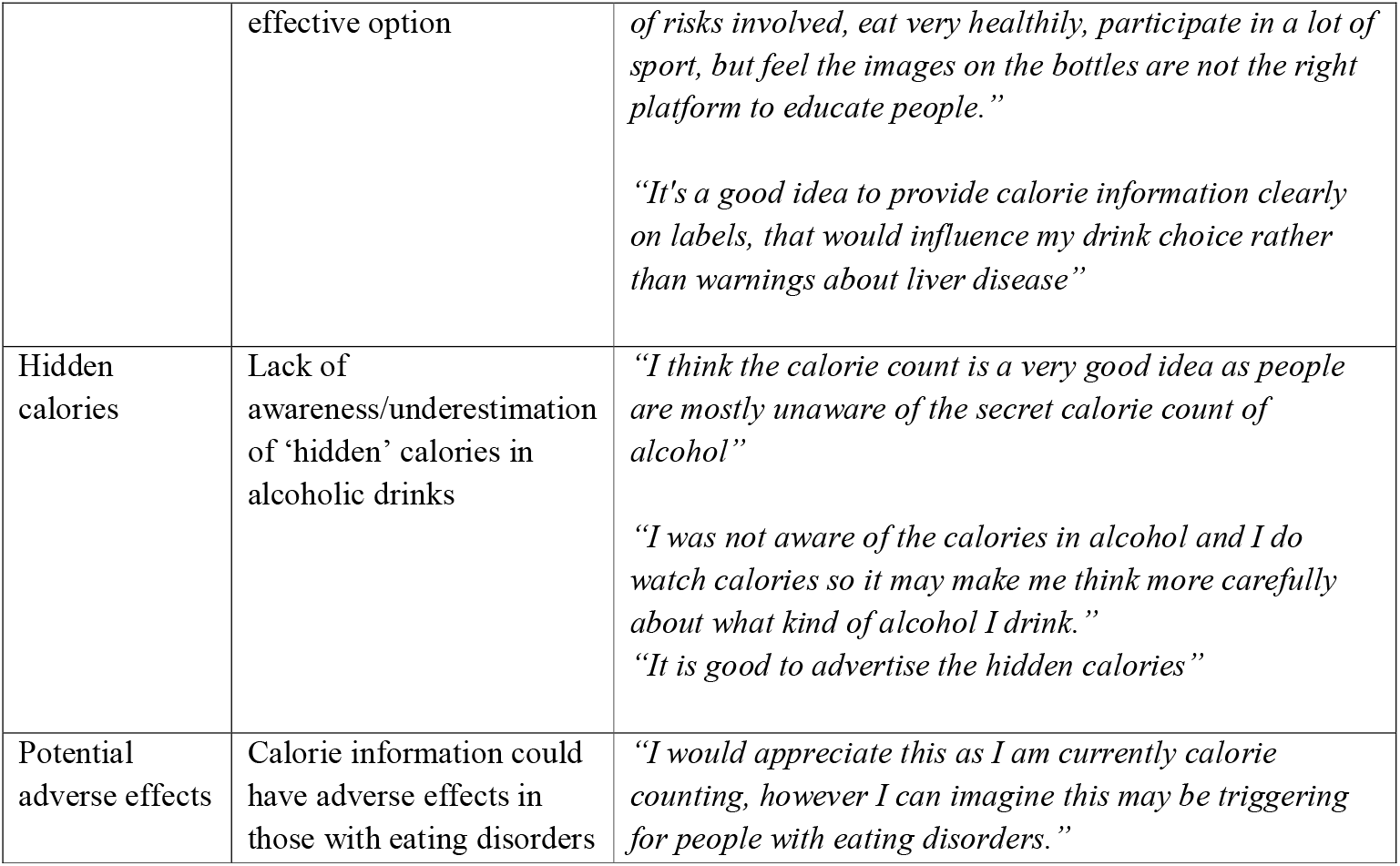
Calorie specific comments.

## Discussion

We found no evidence that either health warning labels – image-and-text or text-only - describing the adverse effects of alcohol consumption, or calorie labels changed the number of alcohol units selected or purchased in an online purchasing setting.

In pre-specified subgroup analyses comparing the ‘calorie label only’ group to the ‘no label’ group, there was no effect on calorie selection. Of those who went on to purchase drinks there was some evidence suggesting that calorie labels on alcoholic and non-alcoholic drinks might reduce calories purchased from both types of drinks.

### Interpretation of findings

The null findings for the impact of HWLs on all selection and purchasing outcomes were contrary to predictions, although these predictions were made in the context of extremely limited pre-existing evidence. The findings accord with our previous experimental study in a naturalistic shopping setting, where the same image-and-text and text-only HWLs as used in the current study also had no impact on selection ^15^. Previous studies that have reported positive effects of alcohol HWLs on selection or purchasing behaviour have predominantly been conducted in online hypothetical settings ^4,10,13,30^, suggesting differences in findings may be explained by the study setting and/or nature of the outcome measure ^14^. It may therefore be that HWLs – in either image-and-text or text-only form - are not sufficient to change real-world purchasing behaviour. Alternatively, it may be that short-term exposure to HWLs on a single shopping occasion is insufficient but longer term or repeated exposure is. The only other study that we are aware of that included actual (i.e., not hypothetical) behavioural outcomes found that labels that included text-only health warnings reduced sales of alcohol over a 14-month period ^31^, suggesting potential longer-term effects. However, this study could not isolate the specific impact of the warning label as there were multiple labels in rotation and the warnings were halted after one month due to industry backlash ^32^.

In a planned analysis comparing only two groups there was no evidence for an effect of calorie labelling on alcohol selection. In those participants who went on to purchase drinks, fewer calories were purchased from both alcoholic and non-alcoholic drinks. Although these results suggest calorie labelling may have the potential to impact purchasing behaviour, due caution should be applied given they were from a subgroup analysis with a relatively small sample size. Furthermore, exploratory analyses suggested that the 77% of participants who went on to purchase differed in their demographic and drinking characteristics - they tended to be older, more educated, and self-reported drinking less alcohol. The only comparable evidence to date on calorie purchasing comes predominantly from food and soft drink studies where a Cochrane review – currently being updated ^33^ – identified limited evidence suggesting small effects on purchasing, but with considerable uncertainty ^16^. There was no evidence for an effect of calorie labels when they were combined with health warning labels. This could be explained by an information overload effect, which posits that too much information on products can overload cognitive capacity and impair the quality of decisions ^34^. Alternatively, it could simply be that HWLs distracted participants from the calorie label.

In terms of acceptability, calorie labels were rated as acceptable, and more so than either type of HWL. This accords with the majority of free-text comments being coded as positive, as well as other evidence of public support for calorie labelling on alcohol ^35^. Calorie labels were also rated lower on negative emotional arousal. Text-only HWLs were rated as higher on acceptability and lower on negative emotional arousal than image-and-text HWLs, consistent with previous studies ^10,15,23^.

Free-text comments indicated calorie information was perceived as a helpful intervention to encourage healthier choices through switching to lower calorie options, as well as highlighting the calories contained in alcohol, of which many people are unaware ^22^. However, previous research has also suggested that calorie labelling could encourage compensatory behaviours with the potential to harm health, such as reducing food intake or selecting drinks with fewer calories but greater alcohol content, to maximise alcohol intake while minimising energy intake ^36^. The possibility of harmful unintended consequences of calorie labelling warrant further study. Attitudes towards HWLs evident in free-text comments in the current study were similar to those observed in previous studies, with negative emotional reactions – including shock, disgust, and fear - being common and mixed views concerning their potential effectiveness and acceptability ^15,23^.

### Strengths and limitations

To our knowledge this is the first randomised controlled trial to estimate the impact of HWLs and calorie labelling on drinks in a naturalistic setting. Meaningful selection and actual purchasing outcomes were assessed, with participants able to complete their typical online shop, including selecting and purchasing freely from a wide range of drinks.

The study also had some limitations. First, while the primary selection outcome was assessed with a stated *intention* to purchase, subsequent purchasing was not mandated or forced, and there was substantial drop-out (23%) between selection and actual purchasing. Although this study required participants to transfer between simulated and actual supermarkets which may have exacerbated the degree of attrition, some drop-out may be inevitable when assessing behaviour in online shopping contexts, given the inevitable time gaps between selecting and ultimately purchasing products. For example, ‘cart abandonment’ – where people do not purchase items they put in their shopping cart – is common in online (including supermarket) shopping contexts ^37^. Retention to the point of actual purchasing was also significantly improved (from 66% to 77%) relative to our previous study using a similar protocol ^38^, likely explained by an increased financial incentive and the refinement of study instructions. Additionally, the majority of participants that did purchase also went on to purchase the exact drinks they selected, indicating this study procedure is feasible and effective in measuring objective selection and purchasing in online shopping settings; future studies using a similar method should account for a similar degree of attrition.

Second, participants in the current study sample were of a higher socioeconomic status than the average in the UK ^39^, although this is likely representative of those who regularly purchase online at Tesco ^40^ and who consume alcohol ^41^. As discussed in the ‘Interpretation of findings’ section, exploratory analyses also suggested that participants that went on to purchase in whom calorie labels did reduce purchasing - differed in their demographic and drinking characteristics. Given previous research suggests certain groups are more likely to use calorie information ^42^, further studies on calorie labels in a wider range of populations are required. Differences between selection and purchasing in the current study could also be explained by participants who dropped out before purchasing never having been intending to purchase and/or being less engaged with the study.

Third, the sample size was determined based on available resource, therefore it may be that some effects were smaller than the study was powered to detect. For example, in the planned subgroup analyses, reductions in calories selected of -5% to -10% were observed that were not statistically significant but which could be potentially meaningful reductions at population-level. Future studies should be suitably powered to detect smaller effects.

### Implications for future research and policy

This study suggests that short-term exposure to HWLs may not be sufficient to change purchasing behaviour. The impact of longer-term or repeated exposure is unknown and merits investigation.

Calorie labels show promise and warrant further evaluation, particularly current government interest in their potential implementation in the UK ^17,20^ and internationally; for example Ireland recently passed legislation that requires energy content information on alcohol packaging ^43^. The World Health Organisation recommends that successful alcohol labelling legislation should include information about the harm from alcohol ^44^ and be consistent with non-alcoholic drink labelling, including the provision of calorie information ^43^. Regardless of whether labelling can elicit meaningful effects on behaviour, information on calories can enable people to accurately estimate calorie intake from drinks ^45^ and appears to be highly acceptable to the public. It may also lead to indirect impacts, for example by encouraging industry and supermarkets to increase the availability or promotion of lower calorie alternatives ^33,46,47^.

## Conclusion

There was no evidence that health warning labels changed the number of alcohol units selected or purchased in an online purchasing setting. There was some evidence suggesting that calorie labels on alcoholic and non-alcoholic drinks might reduce calories purchased from both types of drinks. Given this is the first study to date assessing the impact of calorie labels on alcohol selection and actual purchasing, considerable caution is needed in interpreting these findings. Further evaluation is warranted in suitable powered studies in real-world settings.

## Data Availability

Data will be available from the Open Science Framework (together with the study protocol and statistical analysis plan, already uploaded) and the University of Cambridge Research Repository upon publication.

https://osf.io/w5xf8/

## Declarations

### Contributors

GJH, NC, TMM, and MRM conceptualized and designed the study. NC coordinated the study and led on data collection and cleaning with JF, EP and MV. MAP led on the statistical analysis. NC, GJH, and MAP drafted the manuscript with all authors providing critical revisions. All authors had full access to all the data in the study and accept responsibility to submit for publication.

## References

1. World Health Organization (WHO). Global status report on alcohol and health 2018. Available from https://www.who.int/publications/i/item/9789241565639 (accessed June 2022).

2. Murray CJL, Aravkin AY, Zheng P, et al. Global burden of 87 risk factors in 204 countries and territories, 1990–2019: a systematic analysis for the Global Burden of Disease Study 2019. The Lancet 2020; 396: 1223–1249.

3. Hollands GJ, Bignardi G, Johnston M, et al. The TIPPME intervention typology for changing environments to change behaviour. Nature Human Behaviour 2017; 1: 0140.

4. Blackwell AKM, Drax K, Attwood AS, et al. Informing drinkers: Can current UK alcohol labels be improved? Drug and alcohol dependence 2018; 192: 163–170.

5. Royal Society for Public Health (RSPH). Labelling the point: towards better alcohol health information 2018. Available from https://www.rsph.org.uk/our-work/policy/drugs/labelling-the-point.html (accessed June 2022).

6. Hammond D. Impact of the graphic Canadian warning labels on adult smoking behaviour. Tobacco Control 2003; 12: 391–395.

7. Hammond D. Health warning messages on tobacco products: A review. Tobacco Control: An International Journal 2011; 20: 327–337.

8. Canadian Cancer Society. Cigarette Package Health Warnings: International Status Report. 2021. Available from https://cancer.ca/en/about-us/media-releases/2021/international-warnings-report-2021 (accessed June 2022).

9. Thrasher JF, Carpenter MJ, Andrews JO, et al. Cigarette warning label policy alternatives and smoking-related health disparities. American Journal of Preventive Medicine 2012; 43: 590–600.

10. Clarke N, Pechey E, Mantzari E, et al. Impact of health warning labels communicating the risk of cancer on alcohol selection: an online experimental study. Addiction 2021; 116: 41–52.

11. Stafford LD, Salmon J. Alcohol health warnings can influence the speed of consumption. Journal of Public Health (Germany) 2017; 25: 147–154.

12. Wigg S, Stafford LD. Health warnings on alcoholic beverages: Perceptions of the health risks and intentions towards alcohol consumption. PLoS ONE 2016; 11(4):e0153027

13. Kokole D, Anderson P, Jané-Llopis E. Nature and Potential Impact of Alcohol Health Warning Labels: A Scoping Review. Nutrients 2021; 13: 3065.

14. Clarke N, Pechey E, Kosīte D, et al. Impact of Health Warning Labels on Selection and Consumption of Food and Alcohol Products: Systematic Review with Meta-analysis. Health Psychology Review 2020; 1–39.

15. Clarke N, Blackwell A, De-loyde K, et al. Health warning labels and alcohol selection: an experiment in a naturalistic shopping laboratory. Addiction 2021. 116(12):3333–3345

16. Crockett RA, King SE, Marteau TM, et al. Nutritional labelling for healthier food or non□alcoholic drink purchasing and consumption. Cochrane Database of Systematic Reviews. 2018.

17. Department of Health and Social Care (DHSC). Calorie labelling in the out of home sector: implementation guidance. 2021. Available from https://www.gov.uk/government/publications/calorie-labelling-in-the-out-of-home-sector/calorie-labelling-in-the-out-of-home-sector-implementation-guidance (accessed June 2022).

18. Alcohol Health Alliance (AHA). Contents unknown: How alcohol labelling still fails consumers. 2022. Available from https://ahauk.org/wp-content/uploads/2022/06/Labelling_Report_2022_.pdf (accessed July 2022).

19. Royal Society for Public Health (RSPH). Increasing awareness of ‘invisible’ calories from alcohol. Available from https://www.rsph.org.uk/static/uploaded/979245d2-7b5d-4693-a9b3fb1b98b68d76.pdf

20. Department of Health and Social Care (DHSC).Tackling obesity: empowering adults and children to live healthier lives. 2020. Available from https://www.gov.uk/government/publications/tackling-obesity-government-strategy/tackling-obesity-empowering-adults-and-children-to-live-healthier-lives (accessed June 2022).

21. Alcohol Health Alliance (AHA). Joint-letter-on-alcohol-labelling-to-Matt-Hancock-1. 2021. Available from https://ahauk.org/wp-content/uploads/2021/05/Joint-letter-on-alcohol-labelling-to-Matt-Hancock-1.pdf (accessed June 2022)

22. Robinson E, Humphreys G, Jones A. Alcohol, calories, and obesity: A rapid systematic review and meta-analysis of consumer knowledge, support, and behavioral effects of energy labeling on alcoholic drinks. Obesity Reviews 2021; 22: e13198.

23. Pechey E, Clarke N, Mantzari E, et al. Image-and-text health warning labels on alcohol and food: potential effectiveness and acceptability. BMC Public Health 2020; 20: 376.

24. Hammond D. Tobacco Labelling & Packaging Toolkit: A guide to FCTC Article 11. 2009. Available from https://tobaccolabels.s3.ca-central-1.amazonaws.com/uploads/2013/12/IUATLD-Tookit-Complete-Mar-3-2009.pdf (accessed June 2022)

25. Kees J, Burton S, Craig Andrews J, et al. Understanding How Graphic Pictorial Warnings Work on Cigarette Packaging. Journal of Public Policy & Marketing 2010, 29(2), 265–276

26. Reynolds JP, Archer S, Pilling M, et al. Public acceptability of nudging and taxing to reduce consumption of alcohol, tobacco, and food: A population-based survey experiment. Social Science & Medicine 2019; 236: 112395.

27. Office for National Statistics (ONS). Primary set of harmonised concepts and questions. 2016. Available from https://webarchive.nationalarchives.gov.uk/ukgwa/20160106185646/http://www.ons.gov.uk/ons/guide-method/harmonisation/primary-set-of-harmonised-concepts-and-questions/index.html (accessed June 2022).

28. NI Direct. Qualifications: what the different levels mean. 2015 https://www.nidirect.gov.uk/articles/qualifications-what-different-levels-mean x(2015, accessed June 2022).

29. Saunders JB, Aasland OG, Babor TF, et al. Development of the Alcohol Use Disorders Identification Test (AUDIT): WHO Collaborative Project on Early Detection of Persons with Harmful Alcohol Consumption--II. Addiction 1993; 88: 791–804.

30. Davies EL, Foxcroft DR, Puljevic C, et al. Global comparisons of responses to alcohol health information labels: A cross sectional study of people who drink alcohol from 29 countries. Addictive Behaviors 2022; 131: 107330.

31. Zhao J, Stockwell T, Vallance K, et al. The Effects of Alcohol Warning Labels on Population Alcohol Consumption: An Interrupted Time Series Analysis of Alcohol Sales in Yukon, Canada. Journal of Studies on Alcohol and Drugs 2020; 81: 225–237.

32. Vallance K, Stockwell T, Hammond D, et al. Testing the Effectiveness of Enhanced Alcohol Warning Labels and Modifications Resulting From Alcohol Industry Interference in Yukon, Canada: Protocol for a Quasi-Experimental Study. JMIR Research Protocols 2020; 9: e16320.

33. Clarke N, Marteau TM, Pilling M, et al. Energy (calorie) labelling for healthier selection and consumption of food or alcohol. Cochrane Database of Systematic Reviews. 2021. Protocol.

34. Sasaki T, Becker DV, Janssen MA, et al. Does greater product information actually inform consumer decisions? The relationship between product information quantity and diversity of consumer decisions. Journal of Economic Psychology 2011; 32: 391–398.

35. Institute of Alcohol Studies (IAS). IAS calorie labelling report. Alcohol energy (calorie) labelling: Evidence, public support, alternatives, and wider labelling considerations. 2021. Available from https://www.ias.org.uk/report/alcohol-energy-calorie-labelling-evidence-public-support-alternatives-and-wider-labelling-considerations/ (accessed June 2022)

36. Maynard OM, Langfield T, Attwood AS, et al. No Impact of Calorie or Unit Information on Ad Libitum Alcohol Consumption. Alcohol and Alcoholism 2018; 53: 12–19.

37. Shopping cart abandonment rate by industry 2021. Statista. Available from https://www.statista.com/statistics/457078/category-cart-abandonment-rate-worldwide/ (accessed June 2022).

38. Clarke N, Blackwell AK, Ferrar J, et al. Impact on alcohol selection and purchasing of increasing the proportion of non-alcoholic versus alcoholic drinks: randomised controlled trial. 2022; MedRxiv preprint: https://doi.org/10.1101/2022.03.04.22271898

39. Kiszko KM, Martinez OD, Abrams C, et al. The influence of calorie labeling on food orders and consumption: A review of the literature. Journal of Community Health 2014; 39: 1248–1269.

40. OECD. Education at a Glance 2021: OECD Indicators. Available from https://www.oecd.org/education/education-at-a-glance/ (accessed June 2022)

41. International Data Group (IDG). Trading with Tesco. 2017. Available from https://shoppervista.igd.com/articles/article-viewer/t/infographic---trading-with-tesco/i/16633 (accessed June 2022)

42. Beard E, Brown J, West R, et al. Associations between socio-economic factors and alcohol consumption: A population survey of adults in England. PLOS ONE 2019; 14: e0209442.

43. Jané-Llopis E, Kokole D, Neufeld M, et al. What is the current alcohol labelling practice in the WHO European region and what are barriers and facilitators to development and implementation of alcohol labelling policy? WHO health evidence network synthesis report 2020. Available from https://www.euro.who.int/en/health-topics/disease-prevention/alcohol-use/publications/2020/what-is-the-current-alcohol-labelling-practice-in-the-who-european-region-and-what-are-barriers-and-facilitators-to-development-and-implementation-of-alcohol-labelling-policy-2020 (accessed June 2022).

44. World Health Organization (WHO). Health warning labels on alcoholic beverages: opportunities for informed and healthier choices. 2022. Available from https://www.who.int/publications-detail-redirect/9789240044449 (accessed July 2022).

45. Hobin E, Weerasinghe A, Schoer N, et al. Efficacy of calorie labelling for alcoholic and non-alcoholic beverages on restaurant menus on noticing information, calorie knowledge, and perceived and actual influence on hypothetical beverage orders: a randomized trial. Canadian Journal of Public Health 2022; 113: 363–373.

46. Grummon AH, Petimar J, Soto MJ, et al. Changes in Calorie Content of Menu Items at Large Chain Restaurants After Implementation of Calorie Labels. JAMA Network Open 2021; 4: e2141353.

47. Robinson E, Marty L, Jones A, et al. Will calorie labels for food and drink served outside the home improve public health? BMJ 2021; 372: n40.

