## Supplementary Material for "Impact of health warning labels and calorie labels on selection and purchasing of alcoholic and non-alcoholic drinks: a randomised controlled trial"

Supplementary S1. Drink options in the selection task and example images	Page 2

Supplementary S2. Model results for primary and secondary outcomes	Page 6

Supplementary S3. Free-text comment analysis	Page 9

Supplementary S4. Demographic characteristics: exploratory analysis	Page 15

Supplementary S5. Additional outcomes	Page 16

Supplementary S6. Label ratings	Page 18

References	Page 19

**Supplementary S1. Drink options in the selection task**

Alcohol-free beer, alcohol-free cider and alcohol-free wine options used in the selection task were selected based on brand and size matches, where possible, with alcohol options available online at Tesco.com. Additional alcoholic beer, cider and wine was selected based on the leading brands of lager, ale, mild and stout,^1^ cider^2^ and wine^3^ in Great Britain according to the number of users. Alcohol-free beer, cider and wine were clearly labelled to ensure that they are not confused with alcoholic drinks. Drink images were shown as bottles or cans, either individually or in multi-packs. Participants were able to select as many drinks as they would like to purchase in their household grocery shop. For practical reasons, non-alcoholic drinks of interest were restricted to products categorised as either alcohol-free beer (≤0.5% alcohol by volume, ABV), alcohol-free cider (≤0.5% ABV), alcohol-free wine (≤0.5% ABV), or soft drinks that are not aimed at children (e.g., premium still or sparkling fruit flavoured drinks, or mixers such as tonic water, soda water or ginger ale). This definition aims to avoid confusion with drinks purchased as part of the grocery shop that may not be considered as alternatives for alcohol as well as drinks for children (e.g., tea, coffee, squash, milk-based drinks, juice).

**Figure S1a. Example drink (image-and-text HWL group)**

(for the full range of labels used see registered protocol: <https://osf.io/ch2sm/>)

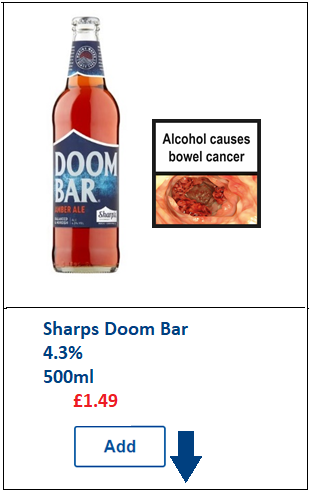

**Figure S1b. Snapshot of simulated store layout (image-and-text HWL group)**

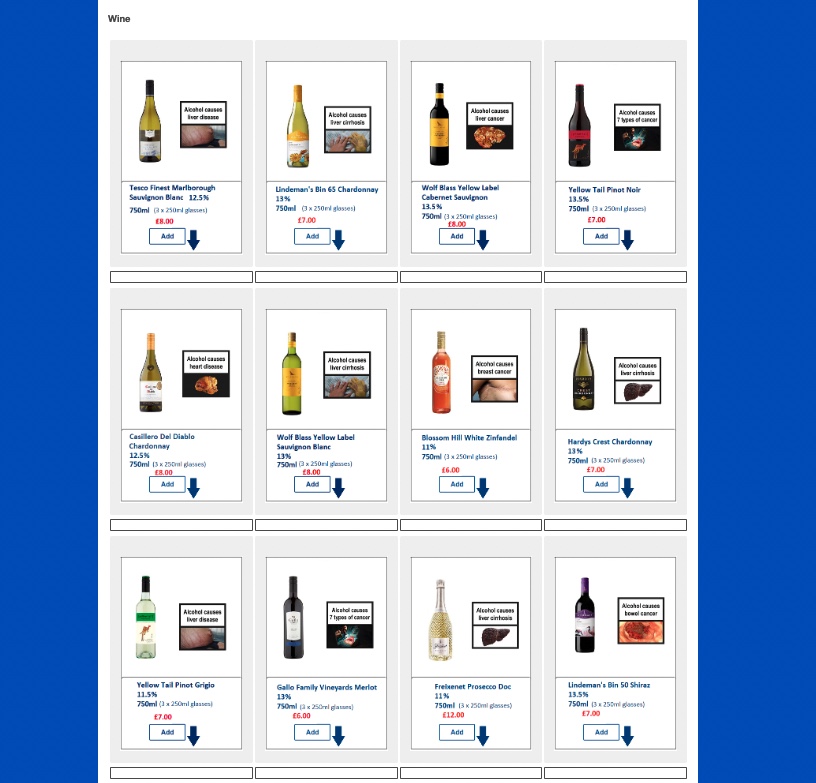

**Table S1. Drink options**

| ***Brand name*** | ***ABV*** | ***Volume*** |
| --- | --- | --- |
| ***Alcohol-free beer and cider*** | | |
| Heineken Alcohol Free Beer | 0.00% | 6 x 330ml |
| Peroni Liberia Alcohol Free Beer | 0.00% | 4 x 330ml |
| San Miguel Alcohol Free Lager | 0.00% | 4 x 330ml |
| Brewdog Punk Nanny State | 0.50% | 4 x 330ml |
| Erdinger Alkoholfrei | 0.50% | 500ml |
| Adnams Ghost Ship Bottle Beer | 0.50% | 500ml |
| Stowford Press Apple Cider Low Alcohol | 0.50% | 500ml |
| Kopparberg Premium Cider Mixed Fruit Alcohol Free | 0.05% | 4 x 330ml |
| ***Alcohol-free wine*** | | |
| Lindeman's Alcohol Free Cabernet Sauvignon | 0.50% | 750ml |
| Tesco Low Alcohol Cabernet Tempranillo | 0.50% | 750ml |
| Eisberg Merlot Alcohol Free Wine | 0.00% | 750ml |
| Hardys Alcohol Free Chardonnay | 0.05% | 750ml |
| Eisberg Sauvignon Blanc Alcohol Free | 0.00% | 750ml |
| Eisberg Rose Alcohol Free | 0.00% | 750ml |
| Freixenet 0.0% Alcohol Free Sparkling | 0.00% | 750ml |
| Nozeco De Alcoholised Wine | 0.50% | 750ml |
| ***Soft drinks*** | | |
| Fentimans Curiosity Cola | n/a | 4 x 275ml |
| San Pellegrino Sparkling Water | n/a | 750ml |
| J20 Orange & Passion Fruit | n/a | 6 x 275ml |
| San Pellegrino Sparkling Limonata | n/a | 6 x 330ml |
| Tesco Soda Water | n/a | 1l |
| Schweppes Tonic Water | n/a | 12 x 150ml |
| Fentimans Traditional Ginger Beer | n/a | 4 x 275ml |
| Belvoir Light Elderflower Presse | n/a | 750ml |
| J2O Spritz Apple Watermelon | n/a | 6 x 275ml |
| Shloer Sparkling White Grape Juice | n/a | 750ml |
| Fentimans Traditional Rose Lemonade | n/a | 750ml |
| San Pellegrino Aranciata Rossa | n/a | 6 x 330ml |
| Oasis Summer Fruit | n/a | 1.5l |
| Schweppes Soda Water | n/a | 1l |
| London Essence Orange & Elderflower Tonic | n/a | 6 x150ml |
| Schweppes Canada Dry Ginger Ale | n/a | 1l |
| ***Beer and cider*** | | |
| Heineken | 5.00% | 12 x 330ml |
| Peroni Nastro Azzurro | 5.10% | 4 x 330ml |
| San Miguel | 5.00% | 4 x 330ml |
| Becks Lager Beer | 4.00% | 20 x 275ml |
| Budweiser | 4.50% | 15 x 440ml |
| Stella Artois Premium Lager | 4.60% | 6 x 330ml |
| Brewdog Punk Ipa | 5.40% | 4 x 330ml |
| Hoegaarden White Beer | 4.90% | 4 x 330ml |
| Adnams Ghost Ship | 4.30% | 500ml |
| Sharps Doom Bar | 4.30% | 500ml |
| Hobgoblin Ipa | 5.30% | 500ml |
| Old Speckled Hen Can | 5.00% | 4 x 500ml |
| Friels Vintage Cider | 7.40% | 4 x 330ml |
| Kopparberg Mixed Fruit Cider | 4.00% | 4 x 330ml |
| Stowford Press Apple Cider 4 | 4.50% | 4 x 440ml |
| Kopparberg Pear | 4.50% | 500ml |
| ***Wine*** | | |
| Tesco Spanish Tempranillo | 12.00% | 750ml |
| Lindeman's Bin 50 Shiraz | 13.50% | 750ml |
| Hardys Varietal Range Merlot | 13.00% | 750ml |
| Yellow Tail Pinot Noir | 13.50% | 750ml |
| Gallo Family Vineyards Merlot | 13.00% | 750ml |
| Wolf Blass Yellow Label Cabernet Sauvignon | 13.50% | 750ml |
| Hardys Crest Chardonnay | 13.00% | 750ml |
| Lindeman's Bin 65 Chardonnay | 13.00% | 750ml |
| Wolf Blass Yellow Label Sauvignon Blanc | 13.00% | 750ml |
| Tesco Finest Marlborough Sauvignon Blanc | 12.50% | 750ml |
| Yellow Tail Pinot Grigio | 11.50% | 750ml |
| Tesco Tempranillo Garnacha Rose | 11.50% | 750ml |
| Blossom Hill White Zinfandel | 11.00% | 750ml |
| Freixenet Prosecco Doc | 11.00% | 750ml |
| Tesco Finest Prosecco Doc | 11.00% | 750ml |
| Casillero Del Diablo Chardonnary | 12.5% | 750ml |

**Supplementary S2. Model results for primary and secondary outcomes**

**Table S2a. Model results regression summaries: model estimates, p values, 95%CIs**

|  | **HWL: image-and-text** | **HWL: text-only** | **Calorie label** |
| --- | --- | --- | --- |
| **Primary outcome** | | | |
| Total alcohol units selected | 0.99, p = .596  95% CI: -2.66, 4.64 | 0.25, p = .893  95% CI: -3.45, 3.96 | 0.52, p = .737  95% CI: -2.50, 3.53 |
| **Secondary outcomes: selection** | | | |
| *Alcohol* | | | |
| Number of alcoholic drinks selected | 0.92, p = .282  95% CI: -0.76, 2.60 | 0.28, p = .751  95% CI: -1.43, 1.98 | 0.37, p = .598  95% CI: -1.01, 1.76 |
| Number of non-alcoholic drinks selected | -0.12, p = .90  95% CI: -2.04, 1.79 | 0.15, p = .878  95% CI: -1.79, 2.09 | 1.60, p = .05  95% CI: 0.03, 3.18 |
| Proportion of alcoholic drinks selected that are alcoholic^1^ | 0.061, p = .678 | 0.0057, p = .969 | -0.15, p = .225 |
| *Calories* | | | |
| Total calories selected^2^ | -- | -- | Calorie label main effect:  0.07, p = .266  Group 3 vs Group 6: -0.049, p = .643  5% reduction, 95% CI: -23%, 17% |
| Calories selected from alcoholic drinks | -- | -- | Calorie label main effect:  0.02, p = .844  Group 3 vs Group 6: -0.105, p = .569  10% reduction, 95% CI: -37%, 30% |
| Calories selected from non-alcoholic drinks^3^ | -- | -- | Calorie label main effect:  0.184, p = .0.021  Group 3 vs Group 6: -0.009, p = 0.947  9% reduction, 95%CI: -28%, 22% |
| **Secondary outcomes: purchasing** | | | |
| *Alcohol* | | | |
| Total alcohol units purchased | 3.08, p = .116  95% CI: -0.76, 6.92 | 1.98, p = .310  95% CI: -1.86, 5.85 | 0.17, p = .917  95% CI: -2.99, 3.32 |
| Proportion of alcoholic drinks purchased that are alcoholic^1^ | 0.14, p = .413 | 0.068, p = .687 | -.13, p = .361 |
| *Calories* | | | |
| Total calories purchased ^2^ | -- | -- | Calorie label main effect:  0.002, p = .976  Group 3 vs Group 6: -0.22, p = .0282*  20% reduction, 95% CI: -35%, -2% |
| Calories purchased from alcoholic drinks^3^ | -- | -- | Calorie label main effect  -0.0005, p = .993  Group 3 vs Group 6: -0.25, p = .0229*  22% reduction, 95% CI: -37%, -3% |
| Calories purchased from non-alcoholic drinks^3^ | -- | -- | Calorie label main effect:  0.033, p =.721  Group 3 vs Group 6: -0.416, p = .0086*  34% reduction, 95% CI: -51%, -10% |

^1^beta-binomial regression used for proportion outcomes (note 95%CIs are available around each group effect compared to no label, but not for the main effects)

^2^ Negative binomial regression – confidence intervals reported as percentages

^3^ Hurdle models (part 2) – truncated negative binomial regression, confidence intervals reported as percentages. Part 1 results reported in Table S2b.

*sig at 0.05 level

**Table S2b. Hurdle model results. Part 1 – binary outcomes (logistic): model estimates, p values**

| **Calorie outcome** | **Calorie label** |
| --- | --- |
| Calories selected from non-alcoholic drinks | Calorie label main effect:  0.12, p = .490  Group 3 vs Group 6:  0.30, p = .313 |
| Calories purchased from alcoholic drinks | Calorie label main effect:  -0.13, p = .693  Group 3 vs Group 6:  -0.51, p = .384 |
| Calories purchased from non-alcoholic drinks | Calorie label main effect:  0.12, p = .532  Group 3 vs Group 6:  0.15, p = .646 |

**Supplementary S3. Free-text comment analysis**

Comments provided by participants were analysed by three study authors (NC, EP, MV).

Comments were first coded as intervention-related or unrelated. Intervention-related comments included those that referred to HWLs or calorie labelling, or related to the experience of seeing the labels during the study. They also included more general comments on drinking behaviour or opinions towards alcohol interventions. Comments were also coded as being either referring to HWLs or calorie labels, if a specific label type was mentioned they were then further coded into positive or negative sentiment.

Themes

Intervention-related comments were thematically analysed following the six steps outlined by Brand and Clarke (2006)^4^. Comments were initially coded two authors (EP, MV) and provisional codes were created. The themes were then refined by grouping codes through a discussion between the two authors and a third author (NC), following which themes and subthemes were generated, reviewed and revised. Themes were then agreed by all authors.

**Table S3: Six themes emerging from a thematic analysis of free-text comments**

| **Theme** | **Subthemes and descriptions** | **Example comments** |
| --- | --- | --- |
| **Potential impact of labelling** | **How people purchase**  Labelling was highlighted as changing purchasing behaviour for some, with suggestions that they would lead to fewer alcoholic drinks being purchased. Others said it did and/or would not impact their purchasing. Some participants highlighted that labels would not work and that approaches such as reducing the price of non-alcoholic options, reducing advertising or education would be more effective.  **How people think**  Labelling might help people think more carefully about drinking behaviour  **How people feel**  Emotional reactions to the labels, such as being off-putting or scary | *“The labels put me off buying the alcoholic drinks”.*  *“The alcohol I have chosen to buy is in line with wat I would normally buy and prefer to drink. The labels did not impact my choice.”*  *“non alcoholic wine, beer and spirits need to be cheaper and taste better in order to incentivise a change in behaviour.”*  *“I understand why the labels might seem necessary but, like cigarette labels, they use scare tactics when actually brands and supermarkets should be looking at educating customers and not offering so many deals”*  *The labels did make me think but did not affect my choice on this occasion*  *Has certainly left me thinking about my alcohol consumption.*  *Yes ...Very off putting*  *the labels were scary* |
| **Acceptability of labelling in general** | **Labelling as acceptable**  Some viewed increasing access to information as a positive step for consumers, increasing knowledge and enabling informed choices.  **Labelling as unacceptable**  Labelling as an unacceptable approach which removes the pleasure from drinking and punishes consumers. | *I think anything that gives the consumer more power and information about what they are buying is a great thing. They can choose what to do after being well informed rather than in the dark.*  *There is nothing wrong with informing our choices*  *The labels are off-putting but it raises awareness*  *I think this is a good idea as people tend to forget about the consequences of drinking and health problems it can cause, I do that's for sure*  *I do not like the labels whatsoever - most people realise the risk and don't appreciate be babied by the government while simultaneously paying outrageous amounts of tax for the pleasure*  *Somewhat takes the pleasure out of choosing a nice bottle of wine you enjoy*  *I think the idea of putting these kind of images on alcohol are very damaging because it is under the assumption that everyone drinks too much and almost punishing people that drink in moderation. I think personally the images go way too far and that a simple warning message in text would suffice as no one wants to do their shopping looking at biological images. Imagine if they did that for everything like food as an example i.e too much meat damages this that and the other. Too many haribo rot your teeth (Cue rotten teeth image or haribo pack).* |
| **Reflections on drinking behaviour in relation to labels** | **Drinking in moderation**  If personal drinking behaviour was not viewed as problematic, then labels were viewed as potentially useful for others but not aimed at them.  **Heavier drinkers**  Labels might work differently depending on consumption level and dependency. | *“The labels didn’t put me off as I am aware of my drinking and know I don’t drink anywhere harmful level.”*  *“The labels are very off putting when ordering drinks as they are not nice to see and they do make you think twice but as I don’t consider myself to be a heavy drinker I wasn’t fully put off by the labels and I would still continue to purchase and consume the items.”*  *“I think the labels will deter some people but unfortunately they will not deter those who are already alcoholics”*  *I think the labels might be useful for people who drink more than the recommended amounts to take a moment and think about their behavior.* |
| **Awareness of information** | **Lack of awareness**  Surprise at the content of the labels, for example at the calorie content or health messages.  **Existing awareness**  Information on the labels is not new, participants are already aware | *Surprised at the proposed drinking warning label.*  *The labels were shocking for some of the items as I never think about the calories in alcoholic drinks like I would other drinks.*  *I didn't know about the connection between alcohol and breast cancer. That made me stop and think*  *The labels I find are quite abrupt and I am already aware of the risk no this would not deter me from drinking however it may help educate some people especially younger people*  *The labels were explicit - didn't tell me much that I didn't know (I knew about bowel, liver cancer from alcohol but not breast cancer) but still took me back a little* |
| **Reflections on health warning labels** | **Negative emotions**  Negative emotional reactions towards HWLs, such as shock, fear and disgust.  **Comparison to tobacco HWLs**  Comparing alcohol HWLs to existing warnings on tobacco. | *Some medical warnings were disturbing and very graphic*  *I did find the warning labels a little disturbing , as , whilst we are now used to seeing them on things such as cigarette packets , it feels like a step too far for alcohol, as , I would imagine , the majority of people who do enjoy alcohol, do so moderately and sensibly , and as a minor leisure activity , eg , a glass of wine with a meal, or a beer in the evening .*  *Like the kind you find on cigarettes, informative, disgusting and off putting.*  *I think it’s good idea for warning labels, they are on cigarette packets so why not add to alcohol aswel. Its a warning to make people aware* |
| **Reflections on calorie labels** | **Healthfulness**  Calorie information can be helpful for making healthier choices  **Switching to lower calorie options**  Drink comparison based on calorie content and select lower calorie options  **Comparing calorie labels with HWLs**  Calorie labels as more acceptable or effective option  **Hidden calories**  Lack of awareness/underestimation of ‘hidden’ calories in alcoholic drinks  **Potential adverse effects**  Calorie labels could have adverse effects in those with eating disorders | *“I felt more drawn to the alcohol free drinks and will be trying to buy these more as I like to look after my body and diet and seeing the calorie content also made me think that they are a better option.”*  *“I think the labels are a great idea. people are becoming more health conscious so being able to see how may calories are in drinks will help us make healthier choices”*  *“The calories next to the item was useful, as you do start to compare items based on the lowest number”*  *“It's interesting to see that eg the alcohol free beer is only 20 calories and that a glass of low alcohol wine is less than half the calories of a normal glass.”*  *“I'm happy to see the calorie content highlighted, if necessary. However, would prefer not to see sad images on something that equals fun time in my life. I am aware of risks involved, eat very healthily, participate in a lot of sport, but feel the images on the bottles are not the right platform to educate people.”*  *“It's a good idea to provide calorie information clearly on labels, that would influence my drink choice rather than warnings about liver disease”*  *“I think the calorie count is a very good idea as people are mostly unaware of the secret calorie count of alcohol”*  *“I was not aware of the calories in alcohol and I do watch calories so it may make me think more carefully about what kind of alcohol i drink.”*  *“It is good to advertise the hidden calories”*  *“I would appreciate this as i am currently calorie counting, however i can imagine this may be triggering for people with eating disorders.”* |

**Supplementary S4. Demographic characteristics: exploratory analysis**

**Table S4. Demographic characteristics of those who did and those who did not go on to purchase**

|  | **Purchased**  **(n=467)** | **Did not purchase (n=141)** | **P value (statistical test)** |
| --- | --- | --- | --- |
| **Alcohol consumption previous week (units)^1a^** **(mean (SD))** | 21.9 (21.4) | 32.1 (30.8) | p < 0.001(Wilcoxon) |
| **Alcohol purchasing previous week (units)^1b^** **(mean (SD))** | 29.9 (25.5) | 37.1 (28.4) | p = 0.003 (Wilcoxon) |
| Age (mean) | 36.8 (10.9) | 31.2 (9.3) | p < 0.001 (Wilcoxon) |
| **BMI (mean (SD))** | 26.2 (5.7) | 26.3 (5.3) | p = 0.7 (Wilcoxon) |
| *Gender: n (%)* | | | |
| Male | 203 (44%) | 66 (47%) | p = 0.515 (Chi Squared) |
| Female | 263 (56%) | 74 (53%) |  |
| *Highest qualification: n (%)* | | | |
| No qualifications | 3 | 1 | p = 0.01 (Fisher) |
| Qualifications at level 1 and below | 2 | 1 |  |
| GCSE / O Level grade A*‐C or vocational level 2 or equivalents | 47 (10) | 28 (20%) |  |
| A levels or vocational level 3 or equivalents | 85 (19) | 31 (20%) |  |
| Higher Education or professional / vocational equivalents | 328 (70) | 78 (56%) |  |
| Other qualification | 2 | 1 |  |

**Supplementary S5. Additional outcomes**

**Table S5a. Raw means (+/-SDs), by group and outcome: Additional outcomes**

|  | **Calorie label** | | | | | |
| --- | --- | --- | --- | --- | --- | --- |
|  | **Present** | | | **Absent** | | |
|  | **Group 1:**  **Image-and-text HWL**  **n = 101** | **Group 2:**  **Text-only HWL**  **n = 92** | **Group 3:**  **No HWL**  **n = 101** | **Group 4:**  **Image-and-text HWL**  **n = 106** | **Group 5:**  **Text-only HWL**  **n = 104** | **Group 6:**  **No label**  **n = 104** |
| Total number of drinks selected | 16.0 (14.2) | 15 (19.8) | 12.6 (17.0 | 12.8 (10.8) | 12.2 (11.5) | 13.5 (12.9) |
| *Purchasing outcomes* | **n = 74** | **n = 71** | **n = 74** | **n = 85** | **n =84** | **n = 79** |
| Total number of drinks purchased | 15.0 (12.4) | 13.8 (12.5) | 9.55 (6.88) | 12.9 (10.6) | 12.6 (11.3) | 12.8 (9.86) |
| Total alcoholic drinks purchased | 8.78 (9.35) | 7.69 (8.19) | 5.28 (5.46) | 7.49 (7.39) | 7.86 (7.84) | 6.77 (5.73) |
| Total non-alcoholic drinks purchased | 6.23 (7.81) | 6.13 (7.99) | 4.27 (4.80) | 5.41 (7.79) | 4.76 (6.68) | 6.06 (8.12) |

**Table S5b. Model results for additional outcomes: ANCOVA: F value, degrees of freedom, p values (note interaction term not included as p> 0.01)**

|  | **HWL (overall) main effect** | **Calorie labelling main effect** |
| --- | --- | --- |
| Total number of drinks selected | F (2, 599) = 0.346, p = .708 | F (1, 599) = 1.979, p = .160 |
| Total number of drinks purchased | F (2, 462) = 2.537, p = .08 | F (1, 462) = 0.001, p = .971 |
| Total alcoholic drinks purchased | F (2, 462) = 3.509, p = .031 | F (1, 462) = 0.024, p = .871 |
| Total non-alcoholic drinks purchased | F (2, 462) = 0.271, p = .763 | F (1, 462) = 0.043, p = .836 |

**Table S5c. Model results for additional outcomes: regression summaries: model estimates, p values, 95%CIs**

|  | **HWL: image-and-text** | **HWL: text-only** | **Calorie label** |
| --- | --- | --- | --- |
| Total number of drinks selected | 0.73, p = .607  95% CI: -2.06, 3.52 | 0.36, p = .801  95% CI: -2.46, 3.20 | 1.94, p = .099  95% CI: -0.36, 4.24 |
| Total number of drinks purchased | 2.37, p = .05  95% CI: -0.002, 4.74 | 1.85, p = .129  95% CI: -0.53, 4.23 | 0.40, p = .691  95% CI: -1.55, 2.35 |
| Total alcoholic drinks purchased | 1.81, p = .03  95% CI: 0.18, 3.43 | 1.67, p = .046  95% CI: 0.034, 3.30 | 0.21, p = .761  95% CI: -1.13, 1.54 |
| Total non-alcoholic drinks purchased | 0.56, p = .498  95% CI: -1.06, 2.19 | 0.19, p = .824  95% CI: -1.45, 1.82 | 0.19, p = .783  95% CI: -1.15, 1.53 |

**Supplementary S6. Label ratings**

**Table S6. Ratings of labels for (a) negative emotional arousal and (b) acceptability by experimental group (Means (+/- SDs) and model results).**

|  | **Group 1:**  **Image-and-text HWL, with calories**  **n = 101** | **Group 2:**  **Text-only HWL, with calories**  **n = 92** | **Group 3:**  **No HWL, calories only**  **n = 101** | **Group 4:**  **Image-and-text HWL, no calories**  **n = 106** | **Group 5:**  **Text-only HWL, no calories**  **n = 104** |
| --- | --- | --- | --- | --- | --- |
| **Negative emotional arousal** | | | | | |
| Means (+/- SDs) [95%CI] | 4.02 (1.70)  [3.72, 4.33] | 3.10 (1.52)  [2.82, 3.38] | 1.86 (1.18)  [1.65, 2.07] | 4.46 (1.51)  [4.20, 4.72] | 3.63 (1.67)  [3.34, 3.93] |
| Regression (with Group 1 as the reference group): model estimate (SE), p values | -- | -0.96 (SE = 0.2), p < 0.001 | -2.22 (SE = 0.2), p < 0.001 | 0.42 (SE = 0.19), p = 0.03 | -0.42 (SE = 0.2), p = 0.04 |
| **Acceptability** | | | | | |
| Means (+/- SDs) [95%CI] | 4.74 (1.62)  [4.45, 5.02] | 4.65 (1.59)  [4.36, 4.95] | 5.34 (1.42)  [5.08, 5.59] | 4.30 (1.73)  [4.00, 4.60] | 4.96 (1.47)  [4.70, 5.22] |
| Regression (with Group 1 as the reference group): model estimate (SE), p values | -- | -0.08 (SE = 0.2), p = 0.68 | 0.57 (SE = 0.2), p = 0.004 | -0.41 (SE = 0.2), p = 0.04 | -0.21 (SE = 0.2), p = .297 |

**References**

1.	Leading brands of beer in United Kingdom 2020. *Statista*, https://www.statista.com/statistics/868499/leading-brands-of-beer-in-the-uk/ (accessed 25 February 2022).

2.	Leading brands of cider in the UK 2020. *Statista*, https://www.statista.com/statistics/317609/leading-brands-of-cider-in-the-uk/ (accessed 25 February 2022).

3.	Leading brands of still wine in the UK 2020. *Statista*, https://www.statista.com/statistics/304150/leading-brands-of-wine-including-sparkling-gb-in-the-uk/ (accessed 25 February 2022).

4.	Braun V, Clarke V. Using thematic analysis in psychology. *Qualitative Research in Psychology* 2006; 3: 77–101.
